# Mental health trajectories of children and adolescents up to five years after the onset of the COVID-19 pandemic: a longitudinal study

**DOI:** 10.64898/2025.11.30.25340982

**Authors:** Viviane Richard, Elsa Lorthe, Roxane Dumont, Nicolas Bovio, Natalia Fernandez, Mayssam Nehme, Rémy P. Barbe, Klara M. Posfay-Barbe, Idris Guessous, Silvia Stringhini, SEROCoV-KIDS study group

## Abstract

**Background:** The COVID-19 pandemic had heterogeneous effects on the mental health of children and adolescents according to individual experiences, with some consequences persisting beyond the lifting of restrictions. We aimed to examine whether the perceived impact of the COVID-19 pandemic was associated with 2022-2025 trajectories of mental health difficulties in children and adolescents, and to identify associated risk and protective factors.

**Methods:** Data was drawn from the population-based SEROCoV-KIDS cohort study conducted in Geneva, Switzerland. The multidimensional perceived impact of the pandemic, as well as potential socio-demographic, health, family, social, and behavioral risk and protective factors were parent-reported at baseline, in 2022. Mental health difficulties were collected annually between 2022 and 2025. Generalized mixed effects models were used to estimate mental health trajectories by pandemic impact, and to assess risk and protective factors.

**Results:** Of 1907 children aged 2-17 years, 9.3% and 7.9% had experienced a negative and positive pandemic impact, respectively. Compared to their unaffected peers, negatively impacted children had more mental health difficulties in 2022 (incidence rate ratio [IRR]: 1.51; 95% confidence interval [CI]: 1.34-1.70) and improving trends between 2022 and 2025 (IRR: 0.97; 95% CI: 0.95-1.01). An average-to-poor financial situation was related to a milder mental health response to a negative impact in 2022 (IRR: 0.64; 95% CI: 0.46-0.89). A positive pandemic impact tended to be associated with higher difficulties in 7-12 years old children only (IRR: 1.36; 95% CI: 0.98-1.89) in 2022, with stable trends over time.

**Conclusion:** About five years after the onset of COVID-19, the lasting mental health difficulties presented by negatively impacted children had largely improved. Although globally reassuring, these findings call for proactive measures to prevent such long-term consequences on youth mental health in the event of future crises.

## BACKGROUND

The COVID-19 pandemic and the public health restrictions implemented to curb its spread affected children and adolescents worldwide [1,2]. School closures, cancellation of extracurricular activities and limitation in gatherings heavily impacted their daily life, while family-level challenges, such as occupational and childcare disruptions or severe infection among relatives, had the potential to spill over into family and child functioning [1,2]. The extent of pandemic-related impacts varied according to children’s circumstances. Stressors, such as disturbed routines, impaired family dynamics, and decline in the financial situation were more common in children with disadvantaged economic backgrounds and unfavorable family environments [3]. Conversely, pre-existing parental psychosocial resources, low financial anxiety and high partner satisfaction were related to positive impacts at the family level including improved well-being and routines [4].

In parallel to the widespread pandemic-related disruptions occurring in 2020, a global increase in mental health problems was observed in youth [5,6], which persisted throughout 2021, despite progressive easing of measures [7–9]. Although mental health has improved since then, levels reported in October 2024 in Germany remained worse than pre-pandemic estimates [10]. Beyond these average trends, significant heterogeneity was observed in youths’ mental health response to the pandemic, reflecting varying experiences [11,12]. Negative impacts of the pandemic, such as disrupted routines, impaired family relationships, and fear for loved ones were strongly associated with poorer life satisfaction and mental health during the spring 2020 lockdown [11,13] and up to two years after the onset of the pandemic [3]. On the other hand, positive impacts, including increased time spent with family or doing recreational activities, were related to better functioning and mental health [11].

The consequences of negative and positive pandemic impacts on children’s mental health could be modified by various factors. Girls and older children were more likely to display internalizing symptoms during the pandemic, while boys presented increased externalizing difficulties [6,14–16]. The role of socio-economic circumstances was unclear. An extensive scoping review of longitudinal research published until December 2021 found that disadvantaged children were at higher risk of decreased mental health during the pandemic [17], which could be explained by greater exposure to pandemic-related stressors and limited financial and social resources to face them [2,3]. On the contrary, a meta-analysis including articles with pre-pandemic measurements published until May 2022 showed that children from higher-income backgrounds experienced greater increases in depression and anxiety symptoms, which was possibly related to higher unmet social expectations because of the sanitary restrictions [6]. Additionally, resources such as family and social support were shown to be positively associated with mental health and could act as protective factors [11,12,18]. Similarly, a generally healthier lifestyle during the pandemic was protective of children’s well-being and mental health [19–21], while a worsening of health behaviors, such as increased screen time and decreased physical activity, was not consistently related to decreased mental health [21,22].

Most of these studies were conducted within the first year of the pandemic and did not assess the medium-term consequences on young people’s mental health. Furthermore, while negative effects were extensively studied [5,6,17], potential positive impacts remain poorly investigated. Finally, within the studies investigating risk and protective factors of mental health beyond the first year of the pandemic, only a few could distinguish between general baseline determinants of mental health and determinants of changes during the pandemic. Among those, Zoellner et al. [23] drew on data from the German COPSY cohort collected at five timepoints between May 2020 and October 2022. They found that male sex, younger age, single parenthood, lower parental education, parental depressive symptoms and higher parental burden due to the pandemic were cross-sectionally related to lower child mental health at baseline, while personal psychosocial resources and family cohesion were positively associated. Younger boys living with a single parent were more likely to experience adverse mental health changes over the follow-up period, whereas increased family cohesion and social support were related to positive changes. These results highlight the importance of longitudinal studies evaluating mental health trajectories of children following the pandemic, as well as associated risk and protective factors.

The goals of the current study were therefore to: 1) examine medium-term trajectories of children’s mental health difficulties in relation to both negative and positive impacts of the COVID-19 pandemic in Geneva, Switzerland, and 2) explore how the association between pandemic impact and mental health varied by socio-demographic, health, family, social, and behavioral factors, both cross-sectionally and longitudinally.

## METHODS

### Study design and setting

Data was drawn from the SEROCoV-KIDS population-based prospective cohort study designed to assess the direct and indirect health-related consequences of the pandemic on children and adolescents in Geneva, Switzerland. Inclusion criteria were 1) to be aged between 6 months and 17 years at enrollment, 2) to live in the canton of Geneva, and 3) to have been (or have a sibling) selected from random samples of the population provided by the government administration, or to be part of a family participating in one of the previous population-based serosurveys conducted by our group [24–27].

Baseline assessment took place between December 2021 and June 2022. One parent or referent adult per family filled out a socio-demographic, health and lifestyle questionnaire for each of their participating children, as well as one for themselves and their household. Annual follow-up surveys repeating questions about health were conducted in May-July 2023, 2024, and 2025 with an average of 16.1 (standard deviation [SD]: 2.3), 29.6 (SD: 2.4), and 39.6 months (SD: 2.2) between baseline and follow-up survey completion, respectively. Additionally, thematic questionnaires were distributed in fall 2022 and spring 2024.

For the current analysis, we selected all participants aged between 2 and 17 years old at enrollment and with at least one available mental health assessment. Three children with a sex reported as other were excluded because models could not be estimated with such a small category. A total of 1907 children were included in the analysis.

### Measures

#### COVID-19 pandemic impact

The impact scale of the COVID-19 Exposure and Family Impact Scales (CEFIS), collected in the fall 2022 thematic questionnaire, was used to retrospectively assess the extent to which the pandemic had affected children’s and families’ functioning, as well as their physical and emotional well-being [28]. Questions about how the pandemic had impacted various aspects of life (e.g. family relationships, sleep, mood) were rated on a 5-point Likert scale ranging from “Made it a lot better” to “Made it a lot worse”, with the addition of a “No change” option compared to the original version [29]. Answers were averaged across the 10 items and categorized into *Positive impact* for scores smaller than or equal to one SD below the sample mean, *Negative impact* for scores greater than or equal to one SD above the sample mean, and *No or minimal impact* otherwise. Well-being items were collected for both children and parents through parent-report and combined with family interactions items to construct child– and household-level impact measures, respectively. The child pandemic impact was used as the main exposure, while the household measure was used in sensitivity analyses.

#### Mental health difficulties

Mental health difficulties were assessed with the French version of the parent-reported Strengths and Difficulties Questionnaire (SDQ) at baseline, in 2022, and in the May-July 2023, 2024 and 2025 follow-up questionnaires [30]. Items were added up according to scoring instructions to calculate an overall score of mental health difficulties (possible range 0-40), regarded as the primary outcome, with higher values indicating increased difficulties. Internalizing and externalizing scores were additionally computed for secondary analyses by adding the emotional and peer problems subscales, and by combining the conduct problems and hyperactivity subscales, respectively [31].

#### Socio-demographic, health, family, social, and behavioral factors

Potential risk and protective factors were parent-reported at baseline, in 2022. Socio-demographic characteristics included age, sex, the highest education level among parents (*university* versus *lower than university*), and perceived household financial situation, considered as *very good* if parents reported being able to save money, *good* if they could meet their needs and cover small unexpected expenses, and *average to poor* if they could just meet their needs or had to rely on external support. Chronic condition was defined as a physical health condition or mental behavioral and neurodevelopmental disorder diagnosed by a healthcare professional and lasting over 6 months. Family and social support comprised the referent parents’ rated relationship with the child categorized into *good* and *less than good* for answers such as rather good, rather poor and poor, the referent parents’ mood (*good* versus *average to poor*), the number of close friends (*several* versus *one or none*) collected from 6 years old, and regular participation in extracurricular activities (*yes* versus *no*) collected from 3 years old. Child average screen time, physical activity and sleep duration were reported in hours per day. Adherence to corresponding recommendations was assessed using age-specific thresholds from the World Health Organization [32,33], or when not available, from the Canadian 24-Hour Movement Guidelines [34].

### Statistical Analysis

The mental health difficulties total score was modelled with generalized mixed effects models following a generalized Poisson distribution, using the R glmmTMB package [35], with the child pandemic impact as main exposure. A three-level model was specified with random effects at the child and household levels. The model was adjusted for confounders by including time fixed effects for age, sex, chronic condition and household financial situation (Additional figure 1). Time since baseline was modeled as a time varying effect with a continuous measure to account for the varying intervals between questionnaire completion across children. 2022-2025 trajectories of mental health according to the pandemic impact were estimated by adding an interaction term between the pandemic impact and time. Sensitivity analyses were conducted by replicating the above-described model with the measure of household pandemic impact, with internalizing and externalizing mental health difficulties, as well as for different age groups to examine age-dependent patterns (2-6, 7-12 and 13-17 years old). The moderating effect of baseline socio-demographic, health, family, social, and behavioral factors in the association between the child pandemic impact and overall mental health difficulties over time was separately tested for each moderator by including a moderator × pandemic impact and a moderator × pandemic impact × time interaction terms. Non-linear time trends were tested by adding a quadratic term but were not included in the final models as they did not meaningfully improve the fit.

**Figure 1.**
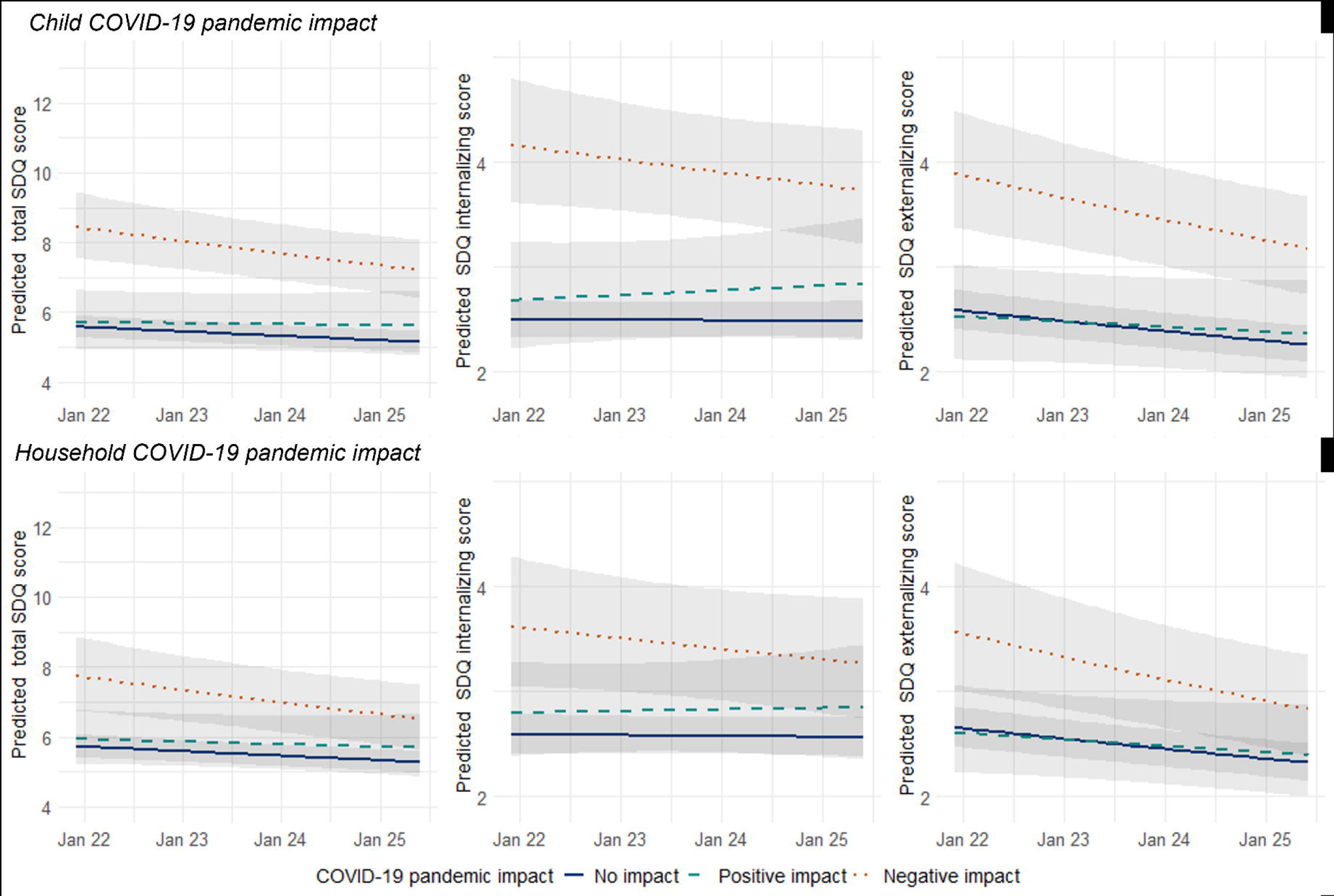
Trajectories of mental health difficulties according to the child and household impact of the COVID-19 pandemic in children aged 2-17 years old, between 2022 and 2025 (n=1907), with higher strengths and difficulties questionnaire (SDQ) scores indicating heightened difficulties. Marginal score prediction and 95% confidence interval (shaded area) from generalized mixed effects models adjusted with fixed effects for age, sex, chronic condition, and household financial situation, and random effects at the child and household levels. Missing values imputed with multiple imputations by chained equations.

Information on the pandemic impact collected in the first thematic questionnaire was available for 1529/1907 (80.2%) children, while mental health data was complete for 1325 (69.5%), 1089 (57.1%), and 971 (50.9%) children at the first, second and third follow-up, respectively. To deal with variable missingness arising from both loss to follow-up and item non-completion (mainly financial situation: 4.0%), multiple imputation was performed. Assuming missingness at random, 40 datasets were imputed by chained equations using the R mice package [36]. The imputation model relied on the above-described variables, as well as on auxiliary variables. This included additional baseline measures of pandemic impact, such as the Coronavirus Impact Scale [37] and questions assessing lasting changes in children’s screen time, physical activity and sleep duration due to the pandemic, as well as one-item measures of perceived physical and mental health collected in each baseline, follow-up and thematic questionnaire. Models based on full information maximum likelihood were estimated as sensitivity analysis.

Analyses were performed with R 4.4.3. GPT-4o was used to improve the readability and language.

## RESULTS

A total of 1907 children from 1134 households were included. There were 952 (49.9%) girls; 419 (22.5%) children were aged 2-6 years old, 873 (47.0%) 7-12 years old, and 566 (30.5%) 13-17 years old (Table 1). According to the measure of child COVID-19 pandemic impact, 9.3% of children had experienced a negative impact and 7.9% a positive impact, while most of them were considered minimally or non-affected (82.8%). A negative impact of the pandemic was more frequently reported in children with a chronic condition, unfavorable family and social situations and not adhering to health behaviors recommendations than in their counterparts (Table 1). An average-to-poor household financial situation was related to both negative and positive impacts.

**Table 1.**
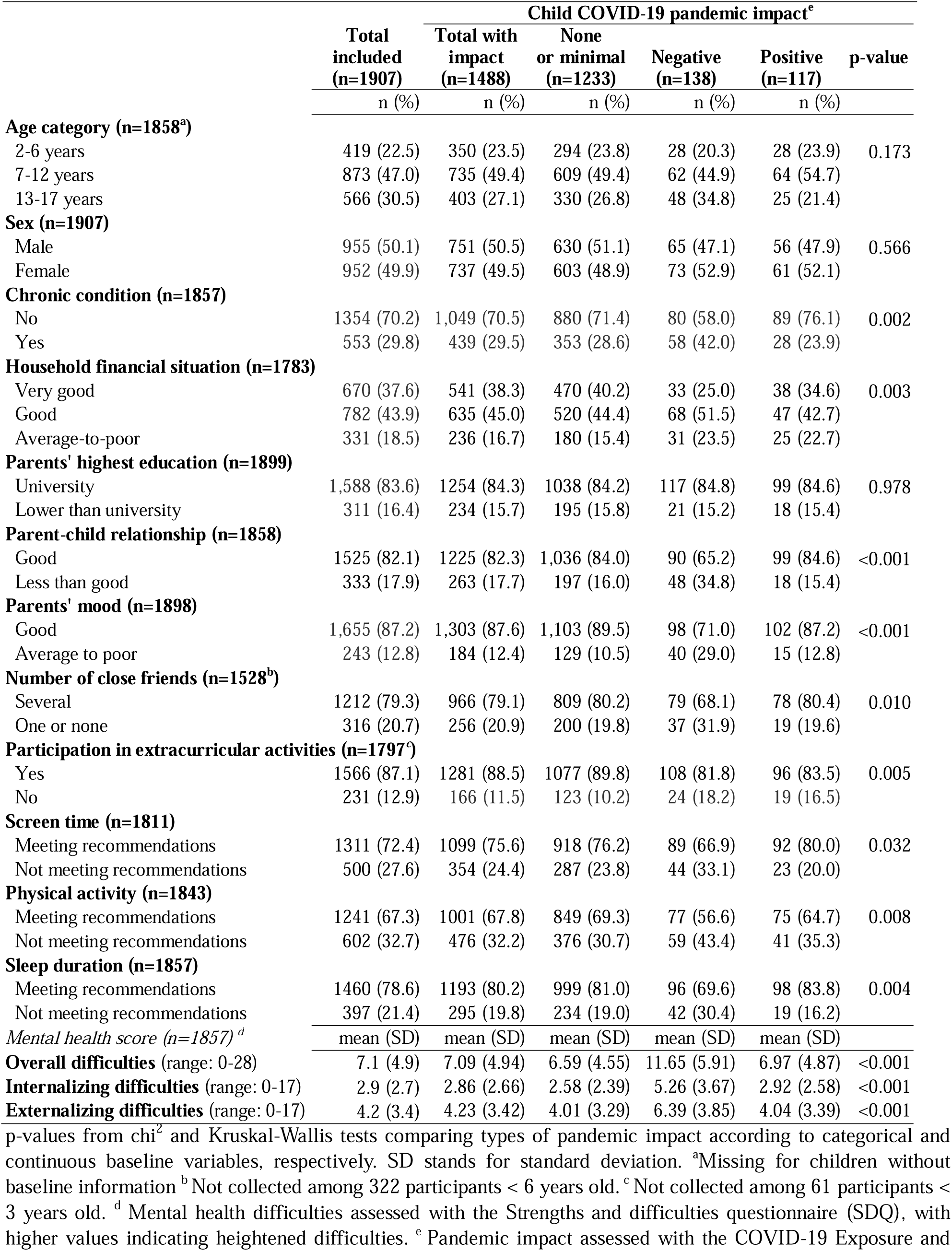
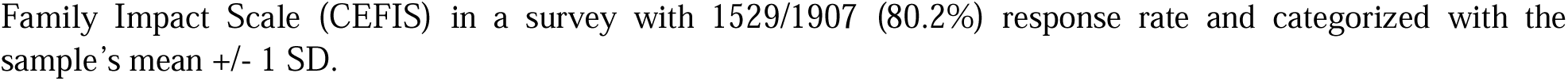
Participants’ baseline characteristics according to the child COVID-19 pandemic impact.

|  | Child COVID-19 pandemic impact<sup>e</sup> |  |  |  |  | p-value |
| --- | --- | --- | --- | --- | --- | --- |
|  | Total included<br>(n=1907) | Total with<br>impact<br>(n=1488) | None<br>or minimal<br>(n=1233) | Negative<br>(n=138) | Positive<br>(n=117) |  |
|  | n (%) | n (%) | n (%) | n (%) | n (%) |  |
| **Age category (n=1858<sup>a</sup>)** |  |  |  |  |  |  |
| 2-6 years | 419 (22.5) | 350 (23.5) | 294 (23.8) | 28 (20.3) | 28 (23.9) | 0.173 |
| 7-12 years | 873 (47.0) | 735 (49.4) | 609 (49.4) | 62 (44.9) | 64 (54.7) |  |
| 13-17 years | 566 (30.5) | 403 (27.1) | 330 (26.8) | 48 (34.8) | 25 (21.4) |  |
| **Sex (n=1907)** |  |  |  |  |  |  |
| Male | 955 (50.1) | 751 (50.5) | 630 (51.1) | 65 (47.1) | 56 (47.9) | 0.566 |
| Female | 952 (49.9) | 737 (49.5) | 603 (48.9) | 73 (52.9) | 61 (52.1) |  |
| **Chronic condition (n=1857)** |  |  |  |  |  |  |
| No | 1354 (70.2) | 1,049 (70.5) | 880 (71.4) | 80 (58.0) | 89 (76.1) | 0.002 |
| Yes | 553 (29.8) | 439 (29.5) | 353 (28.6) | 58 (42.0) | 28 (23.9) |  |
| **Household financial situation (n=1783)** |  |  |  |  |  |  |
| Very good | 670 (37.6) | 541 (38.3) | 470 (40.2) | 33 (25.0) | 38 (34.6) | 0.003 |
| Good | 782 (43.9) | 635 (45.0) | 520 (44.4) | 68 (51.5) | 47 (42.7) |  |
| Average-to-poor | 331 (18.5) | 236 (16.7) | 180 (15.4) | 31 (23.5) | 25 (22.7) |  |
| **Parents' highest education (n=1899)** |  |  |  |  |  |  |
| University | 1,588 (83.6) | 1254 (84.3) | 1038 (84.2) | 117 (84.8) | 99 (84.6) | 0.978 |
| Lower than university | 311 (16.4) | 234 (15.7) | 195 (15.8) | 21 (15.2) | 18 (15.4) |  |
| **Parent-child relationship (n=1858)** |  |  |  |  |  |  |
| Good | 1525 (82.1) | 1225 (82.3) | 1,036 (84.0) | 90 (65.2) | 99 (84.6) | <0.001 |
| Less than good | 333 (17.9) | 263 (17.7) | 197 (16.0) | 48 (34.8) | 18 (15.4) |  |
| **Parents' mood (n=1898)** |  |  |  |  |  |  |
| Good | 1,655 (87.2) | 1,303 (87.6) | 1,103 (89.5) | 98 (71.0) | 102 (87.2) | <0.001 |
| Average to poor | 243 (12.8) | 184 (12.4) | 129 (10.5) | 40 (29.0) | 15 (12.8) |  |
| **Number of close friends (n=1528<sup>b</sup>)** |  |  |  |  |  |  |
| Several | 1212 (79.3) | 966 (79.1) | 809 (80.2) | 79 (68.1) | 78 (80.4) | 0.010 |
| One or none | 316 (20.7) | 256 (20.9) | 200 (19.8) | 37 (31.9) | 19 (19.6) |  |
| **Participation in extracurricular activities (n=1797<sup>c</sup>)** |  |  |  |  |  |  |
| Yes | 1566 (87.1) | 1281 (88.5) | 1077 (89.8) | 108 (81.8) | 96 (83.5) | 0.005 |
| No | 231 (12.9) | 166 (11.5) | 123 (10.2) | 24 (18.2) | 19 (16.5) |  |
| **Screen time (n=1811)** |  |  |  |  |  |  |
| Meeting recommendations | 1311 (72.4) | 1099 (75.6) | 918 (76.2) | 89 (66.9) | 92 (80.0) | 0.032 |
| Not meeting recommendations | 500 (27.6) | 354 (24.4) | 287 (23.8) | 44 (33.1) | 23 (20.0) |  |
| **Physical activity (n=1843)** |  |  |  |  |  |  |
| Meeting recommendations | 1241 (67.3) | 1001 (67.8) | 849 (69.3) | 77 (56.6) | 75 (64.7) | 0.008 |
| Not meeting recommendations | 602 (32.7) | 476 (32.2) | 376 (30.7) | 59 (43.4) | 41 (35.3) |  |
| **Sleep duration (n=1857)** |  |  |  |  |  |  |
| Meeting recommendations | 1460 (78.6) | 1193 (80.2) | 999 (81.0) | 96 (69.6) | 98 (83.8) | 0.004 |
| Not meeting recommendations | 397 (21.4) | 295 (19.8) | 234 (19.0) | 42 (30.4) | 19 (16.2) |  |
| **Mental health score (n=1857)<sup>d</sup>** | mean (SD) | mean (SD) | mean (SD) | mean (SD) | mean (SD) |  |
| **Overall difficulties** (range: 0-28) | 7.1 (4.9) | 7.09 (4.94) | 6.59 (4.55) | 11.65 (5.91) | 6.97 (4.87) | <0.001 |
| **Internalizing difficulties** (range: 0-17) | 2.9 (2.7) | 2.86 (2.66) | 2.58 (2.39) | 5.26 (3.67) | 2.92 (2.58) | <0.001 |
| **Externalizing difficulties** (range: 0-17) | 4.2 (3.4) | 4.23 (3.42) | 4.01 (3.29) | 6.39 (3.85) | 4.04 (3.39) | <0.001 |

p-values from $\chi^2$ and Kruskal-Wallis tests comparing types of pandemic impact according to categorical and continuous baseline variables, respectively. SD stands for standard deviation. <sup>a</sup>Missing for children without baseline information <sup>b</sup>Not collected among 322 participants < 6 years old. <sup>c</sup>Not collected among 61 participants < 3 years old. <sup>d</sup> Mental health difficulties assessed with the Strengths and difficulties questionnaire (SDQ), with higher values indicating heightened difficulties. <sup>e</sup> Pandemic impact assessed with the COVID-19 Exposure and
Family Impact Scale (CEFIS) in a survey with 1529/1907 (80.2%) response rate and categorized with the sample's mean ± 1 SD.

Children aged 13-17 years at baseline, with poorer socio-economic circumstances and lower adherence to health behavior recommendations were more likely to have missing data, although patterns of non-response varied across questionnaires (Additional table 1). Modelling results presented hereafter rely on analysis with imputed data for missing baseline variables and incomplete later questionnaires.

### Mental health difficulties according to the pandemic impact at baseline and over time

At baseline, in 2022, participants who had experienced a negative pandemic impact had 51% higher scores of mental health difficulties (incidence rate ratio [IRR]: 1.51; 95% confidence interval [CI]: 1.34-1.70) than those experiencing no or minimal impact, resulting in a score difference of 3.1 points (Figure 1). There was no significant mental health difference between experiencing a positive and no pandemic impact (p-value > 0.1).

Compared to children with no reported impact, those negatively impacted by the pandemic showed a non-significant annual decrease of 3% (IRR: 0.97; 95% CI: 0.95-1.01) in the overall mental health difficulties score between 2022 and 2025 (Figure 1 & Additional table 2). However, when restricting the analysis to the period between 2022 and 2024, this downward trend became steeper and statistically significant (IRR: 0.96; 95% CI: 0.92-0.99), as mental health difficulties slightly increased again in 2025 among negatively impacted children (Additional figure 2). The evolution of mental health did not differ between not impacted and positively impacted children (Figure 1 & Additional table 2). Similar trends were observed when relying on the pandemic impact measured at the household level (Figure 1). Furthermore, these patterns were consistent across internalizing and externalizing mental health difficulties and across age groups, although not significantly (Figure 1 & Additional table 2).

**Figure 2.**
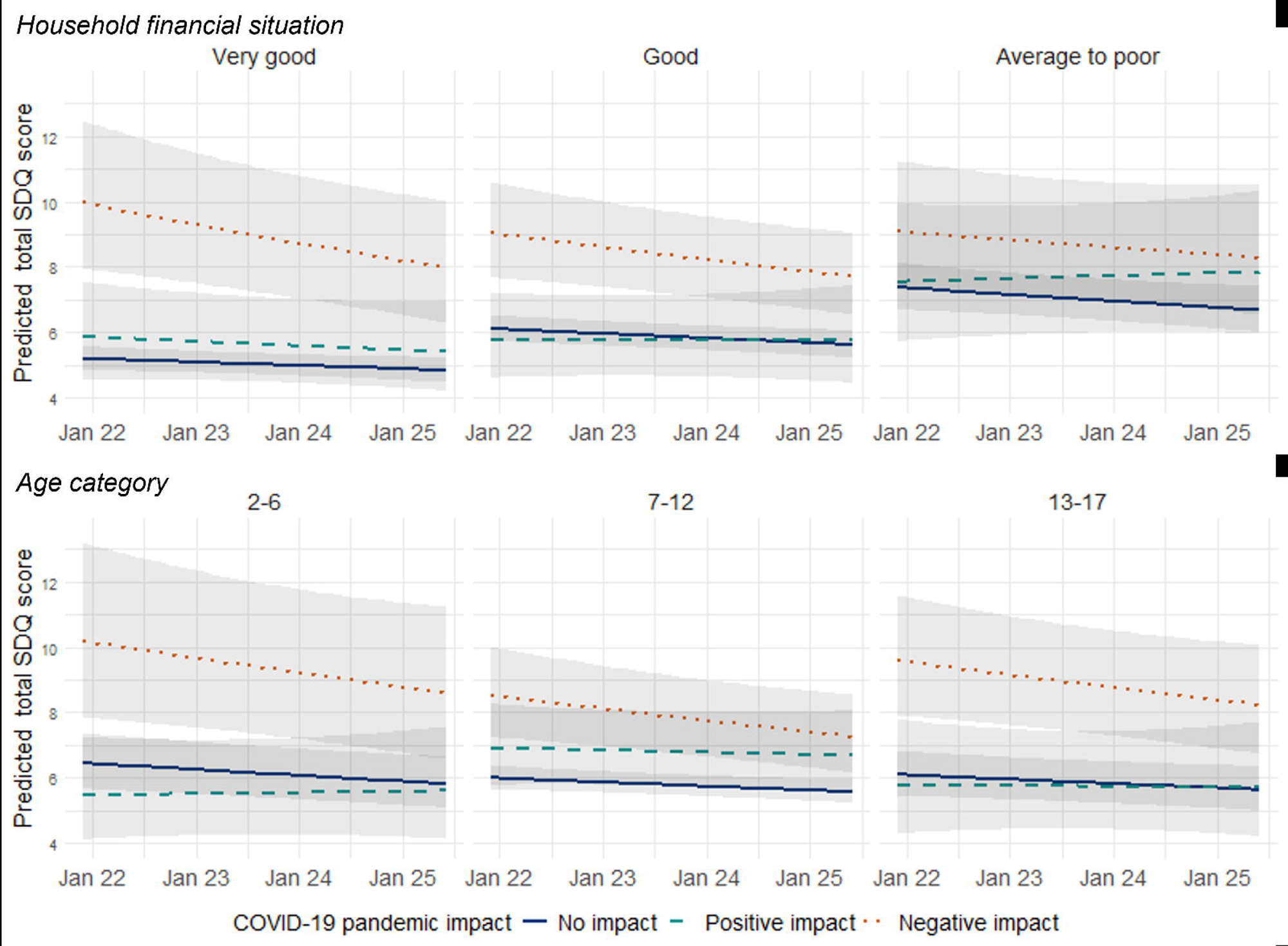
Trajectories of mental health difficulties according to the child impact of the COVID-19 pandemic, household financial situation and age category in children aged 2-17 years old, between 2022 and 2025 (n=1907), with higher strengths and difficulties questionnaire (SDQ) scores indicating heightened difficulties. Marginal score prediction and 95% confidence interval (shaded area) from generalized mixed effects model adjusted with fixed effects for age, sex, chronic condition, and household financial situation, and random effects at the child and household levels. Missing values imputed with multiple imputations by chained equations.

### Risk and protective factors at baseline and over time

When exploring potential moderators, the baseline association between a negative pandemic impact and elevated mental health difficulties was weaker in children with an average-to-poor (IRR: 0.64; 95% CI: 0.46-0.89) or good financial situation (IRR: 0.77; 95% CI: 0.57-1.04) than in their most privileged counterparts (Figure 2 & Additional table 3). However, living in a household with an average-to-poor financial situation (IRR: 1.42; 95% CI: 1.26-1.60), and to a lesser extent a good financial situation (IRR: 1.18; 95% CI: 1.07-1.29), was related to higher average levels of mental health difficulties (Figure 2). A positive pandemic impact tended to be associated with higher baseline mental health difficulties in 7-12 years old children than in younger ones (IRR: 1.36; 95% CI: 0.98-1.89; Figure 2, Additional table 3). There was no other moderator affecting the baseline pandemic impact on mental health difficulties (Additional table 3).

Over time, the rate of change in mental health difficulties according to the pandemic impact did not vary according to any moderator under study (Additional table 3). Analyses with maximum likelihood revealed similar patterns and effect sizes as those conducted with imputed data (Additional table 2 and 3).

## DISCUSSION

In 2022, two years after the onset of COVID-19, children who had experienced a negative impact of the pandemic had higher mental health difficulties than their non-affected counterparts, particularly those with a privileged financial situation. From 2022 to 2024, mental health difficulties largely subsided in negatively impacted children, before slightly increasing again in 2025. Positively impacted 7-12 years old children had more baseline mental health difficulties than their non-impacted peers and followed a stable trend over time, while there was no difference in younger or older children.

Four out of five children were not or only minimally impacted by the pandemic and displayed low levels of mental health difficulties over the 2022-2025 period. It echoes evidence from Germany, where two-third to three-quarter of children followed a stable low trajectory of mental health problems between 2020 and 2022 and constituted the group least burdened by the pandemic [12]. Conversely and as previously observed [11,13], children who were negatively impacted by the pandemic displayed poorer mental health than their unaffected counterparts. Reflecting previous findings [3,10,14], this association was noticeable two years into the pandemic, even after most restrictions had been lifted. We estimated the difference to be of 3.1 SDQ points early 2022, with each additional point corresponding to a 1.28-fold increase in the odds of presenting a mental health disorder [38], which probably constituted an already improved situation compared to the first year of the pandemic [10]. Reassuringly, by summer 2024, the mental health of negatively impacted children had improved and was close to the levels displayed by non-affected children. This is in line with findings from the German COPSY study, showing that the prevalence of mental health difficulties decreased since 2020 but was still slightly higher in October 2024 than before the pandemic [10]. However, a slight rebound in difficulties was observed among negatively impacted children in summer 2025. This trend is unlikely to be attributable to the COVID-19 pandemic, given the absence of an active outbreak and the lifting of pandemic-related measures since February 2022 [39]. However, these children may be particularly sensitive to external stressors and could have been affected by the increasing global economic uncertainty in 2025 [40]. This was a particular concern in Geneva, which hosts many international organizations facing severe budget cuts [41].

An unfavorable financial situation was associated with poorer levels of mental health on average but partly buffered against the effect of a negative pandemic impact on mental health. The observed social gradient in mental health was in line with the extensive literature on health inequalities [42], while pandemic-related patterns echoed meta-analysis and Swiss reports of children with higher socio-economic circumstances experiencing a greater decrease in well-being during the pandemic [6,43]. A potential explanation is that a negative pandemic impact was perceived as a comparatively lesser burden by disadvantaged children who are more familiar with adverse circumstances and could have developed coping strategies to deal with them, than by advantaged children who might have had more unmet expectations [6,8,44]. It could also be that a negative pandemic impact was related to a certain level of heightened mental health difficulties, similar across all children, that represented a greater increase for privileged individuals who had better average levels than their less advantaged counterparts.

A positive pandemic impact tended to be persistently associated with elevated mental health difficulties in 7-12-years old children but not in younger children and adolescents. It may be that school-aged children who enjoyed the lockdown period found it difficult to return to a normal routine after experiencing an improved situation [44]. For instance, this could concern children with learning disorders or social anxiety, who may have benefited from following their own rhythm and being away from daily stressors, including onsite assignment and peer interactions [45]. The onset of such conditions typically occurs from the start of school [46,47], which could explain why a positive pandemic impact was associated with elevated problems in 7-12 but not 2-6-year-old children. In positively impacted adolescents, potential difficulties related to the return to normalcy might have been outweighed by their developmental need for interactions and experiences outside of the family environment [48], hence the absence of difference in this age group.

Even if the improving trend is reassuring, children with the heaviest pandemic burden faced high levels of mental health difficulties up to five years after the onset of the pandemic and may represent a group particularly vulnerable to other major societal events. This highlights the need to consider the psychological well-being of young populations in public responses to crises. Specifically, a consistent routine and age-adapted parent-child communication were shown to be effective in buffering the impact of the pandemic and could be applied to other situations [49]. Since such mitigation approaches rely on the involvement and availability of parents, tailored measures supporting them should also be considered, including flexible working arrangements, when possible, psychological counselling, and financial aid. Finally, policies drawing on the positive aspects of the pandemic could be designed to best fit all children’s needs. For instance, possibilities of adapted education could be explored for children who benefited more from distance than onsite learning [44].

These findings should be interpreted in light of their limitations. First, despite the random selection of participants, children from highly educated parents were more likely to participate, resulting in a sample overrepresenting socially advantaged individuals in comparison to the Geneva population [50]. Therefore, results might not be generalizable to children living in precarious circumstances and might underestimate the association between socio-economic conditions and mental health. Second, the measure of pandemic impact was parent-reported two years into the pandemic and was thus subject to recall and proxy biases, with the perception of the impact severity potentially varying according to personal traits [51]. Third, despite continuous efforts to retain participants, attrition significantly increased over time. However, evidence suggests that approaches such as multiple imputations and full-information maximum likelihood used in the current analysis are robust to loss to follow-up in cohort studies [52]. Finally, we acknowledge that mental health is shaped by a complex and dynamic interplay of multiple determinants, making it challenging to disentangle the effect of the pandemic from age and other factors related to children’s development. Our study also has major strengths including the sizeable population-based sample covering a wide age range, the longitudinal design with multiple assessments up to five years after the COVID-19 pandemic onset, the exploration of the effect of a positive pandemic impact and the inclusion of various socio-demographic, health, family, social, and behavioral moderators.

## CONCLUSION

About five years after the onset of the COVID-19, the lasting mental health difficulties presented by negatively impacted children had largely improved. However, these reassuring findings should not mask that youth mental health was affected for years following the pandemic and might be sensitive to other societal events. Drawing on the extensive research conducted during the pandemic, more consideration and resources should be allocated to foster children’s and adolescents’ mental health in the event of future crises.

## DECLARATION

### Ethics approval and consent to participate

The Geneva Cantonal Commission for Research Ethics approved the study (ID: 2021-01973). All referent adults, as well as adolescents aged 14 years or older provided written consent to participate. Children gave oral assent to participate.

### Consent for publication

Not applicable

### Availability of data and materials

The dataset used during the current study is available from the corresponding author on reasonable request.

### Authors’ contributions

All authors contributed to the study conception and design. Material preparation and data collection were performed by Viviane Richard, Roxane Dumont, Elsa Lorthe, Nicolas Bovio, Natalia Fernandez, Julien Lamour, and Silvia Stringhini. Rémy P. Barbe, Klara M. Posfay-Barbe, Idris Guessous and Silvia Stringhini supervised the study. Analyses were performed by Viviane Richard who also wrote the first draft of the manuscript. All authors critically revised the previous versions of the manuscript. All authors read and approved the final manuscript.

## Data Availability

All data produced in the present study are available upon reasonable request to the authors.

## LIST OF ABBREVIATIONS

CEFIS: COVID-19 Exposure and Family Impact Scales
CI: Confidence Interval
COVID-19: Coronavirus Disease 2019
IRR: Incidence Rate Ratio
SD: Standard Deviation
SDQ: Strengths and Difficulties Questionnaire

## Acknowledgement

We are grateful to the staff of the Unit of Population Epidemiology of the Division of Primary Care Medicine of the University Hospitals of Geneva, as well as to all participants whose contributions were invaluable to the study.

SEROCoV-KIDS study group: Andrew S. Azman, Antoine Bal, Rémy P. Barbe, Hélène Baysson, Aminata R. Bouhet, Nicolas Bovio, Paola D’Ippolito, Roxane Dumont, Nacira El Merjani, Natalia Fernandez Clares, Natalie Francioli, Idris Guessous, Séverine Harnal, Julien Lamour, Arnaud G L’Huillier, Andrea Loizeau, Elsa Lorthe, Chantal Martinez, Shannon Mechoullam, Mayssam Nehme, Klara M. Posfay-Barbe, Géraldine Poulain, Caroline Pugin, Nick Pullen, Viviane Richard, Deborah Rochat, Khadija Samir, Stephanie Schrempft, Silvia Stringhini, Stéphanie Testini, Deborah Urrutia Rivas, Anshu Uppal, Charlotte Verolet, Jennifer Villers, Guillemette Violot, María-Eugenia Zaballa

