## Additional for "Mental health trajectories of children and adolescents up to five years after the onset of the COVID-19 pandemic: a longitudinal study"

Additional files

Viviane Richard, Elsa Lorthe, Roxane Dumont, Nicolas Bovio, Natalia Fernandez, Mayssam Nehme, Rémy P. Barbe, Klara M. Posfay-Barbe, Idris Guessous, Silvia Stringhini, SEROCoV-KIDS study group

**Correspondence to**

Silvia Stringhini

Unité d’épidémiologie populationnelle

Rue Jean-Violette 29

1205 Genève

Switzerland

+41 22 305 58 61

**Additional table 1.** Participants’ baseline characteristics according to response to follow-up surveys.

|  | **Total**  **(n=1907)** | **Survey non-response** | | | | | | | |
| --- | --- | --- | --- | --- | --- | --- | --- | --- | --- |
|  |  | **Thematic survey (n=378)** | **p-value** | **1^st^**  **follow-up (n=585)** | **p-value** | **2^nd^**  **follow-up**  **(n=820)** | **p-value** | **3^rd^ follow-up**  **(n=936)** | **p-value** |
| **Age category (n=1858)** |  |  |  |  |  |  |  |  |  |
| 2-6 years | 419 (22.6) | 54 (16.4) | <0.001 | 126 (22.2) | 0.277 | 163 (20.5) | <0.001 | 183 (20.3) | <0.001 |
| 7-12 years | 873 (47.0) | 120 (36.5) |  | 254 (44.8) |  | 349 (43.9) |  | 397 (44.0) |  |
| 13-17 years | 566 (30.5) | 155 (47.1) |  | 187 (33.0) |  | 283 (35.6) |  | 322 (35.7) |  |
| **Sex (n=1907)** |  |  |  |  |  |  |  |  |  |
| Male | 955 (50.1) | 185 (48.9) | 0.663 | 303 (51.8) | 0.343 | 420 (51.2) | 0.413 | 483 (51.6) | 0.207 |
| Female | 952 (49.9) | 193 (51.1) |  | 282 (48.2) |  | 400 (48.8) |  | 453 (48.4) |  |
| **Chronic condition (n=1857)** |  |  |  |  |  |  |  |  |  |
| No | 1304 (70.2) | 225 (68.6) | 0.521 | 410 (72.3) | 0.211 | 540 (67.9) | 0.069 | 621 (68.8) | 0.227 |
| Yes | 553 (29.8) | 103 (31.4) |  | 157 (27.7) |  | 255 (32.1) |  | 281 (31.2) |  |
| **Household financial situation (n=1783)** | |  |  |  |  |  |  |  |  |
| Very good | 670 (37.6) | 115 (34.3) | 0.003 | 199 (37.5) | 0.338 | 269 (36.0) | 0.372 | 285 (33.1) | <0.001 |
| Good | 782 (43.9) | 136 (40.6) |  | 223 (42.0) |  | 331 (44.3) |  | 396 (45.9) |  |
| Average-to-poor | 331 (18.6) | 84 (25.1) |  | 109 (20.5) |  | 148 (19.8) |  | 181 (21.0) |  |
| **Parents' highest education (n=1899)** |  |  |  |  |  |  |  |  |  |
| University | 1588 (83.6) | 308 (83.2) | 0.887 | 481 (82.4) | 0.357 | 660 (80.9) | 0.006 | 746 (80.3) | <0.001 |
| Lower than university | 311 (16.4) | 62 (16.8) |  | 103 (17.6) |  | 156 (19.1) |  | 183 (19.7) |  |
| **Parent-child relationship (n=1858)** |  |  |  |  |  |  |  |  |  |
| Good | 1525 (82.1) | 266 (80.9) | 0.575 | 466 (82.2) | 0.987 | 640 (80.5) | 0.142 | 744 (82.5) | 0.702 |
| Less than good | 333 (17.9) | 63 (19.1) |  | 101 (17.8) |  | 155 (19.5) |  | 158 (17.5) |  |
| **Parents' mood (n=1898)** |  |  |  |  |  |  |  |  |  |
| Good | 1655 (87.2) | 315 (85.1) | 0.216 | 507 (87.0) | 0.898 | 706 (86.5) | 0.485 | 801 (86.2) | 0.239 |
| Average to poor | 243 (12.8) | 55 (14.9) |  | 76 (13.0) |  | 110 (13.5) |  | 128 (13.8) |  |
| **Number of close friends (n=1528)** |  |  |  |  |  |  |  |  |  |
| Several | 1212 (79.3) | 225 (80.1) | 0.793 | 375 (80.5) | 0.504 | 525 (78.8) | 0.724 | 594 (78.4) | 0.394 |
| One or less | 316 (20.7) | 56 (19.9) |  | 91 (19.5) |  | 141 (21.2) |  | 164 (21.6) |  |
| **Participation in extracurricular activities (n=1797)^a^** |  |  |  |  |  |  |  |  |  |
| Yes | 1566 (87.1) | 264 (82.8) | 0.013 | 469 (85.0) | 0.078 | 645 (83.7) | <0.001 | 745 (84.9) | 0.008 |
| No | 231 (12.9) | 55 (17.2) |  | 83 (15.0) |  | 126 (16.3) |  | 132 (15.1) |  |
| **Screen time (n=1811)** |  |  |  |  |  |  |  |  |  |
| Meeting recommendations | 1311 (72.4) | 184 (57.9) | <0.001 | 389 (71.2) | 0.510 | 521 (67.9) | <0.001 | 590 (67.6) | <0.001 |
| Not meeting recommendations | 500 (27.6) | 134 (42.1) |  | 157 (28.8) |  | 246 (32.1) |  | 283 (32.4) |  |
| **Physical activity (n=1843)^b^** |  |  |  |  |  |  |  |  |  |
| Meeting recommendations | 1241 (67.3) | 214 (65.6) | 0.514 | 374 (67.8) | 0.845 | 519 (66.3) | 0.437 | 590 (66.3) | 0.382 |
| Not meeting recommendations | 602 (32.7) | 112 (34.4) |  | 178 (32.2) |  | 264 (33.7) |  | 300 (33.7) |  |
| **Sleep duration (n=1857)** |  |  |  |  |  |  |  |  |  |
| Meeting recommendations | 1460 (78.6) | 238 (72.3) | 0.003 | 432 (76.2) | 0.103 | 594 (74.7) | <0.001 | 679 (75.3) | 0.001 |
| Not meeting recommendations | 397 (21.4) | 91 (27.7) |  | 135 (23.8) |  | 201 (25.3) |  | 223 (24.7) |  |
| **Mental health difficulties total score^c^** | 7.1 (4.9) | 7.1 (4.9) | 0.955 | 7.2 (4.8) | 0.813 | 7.4 (4.9) | 0.035 | 7.3 (4.8) | 0.210 |
| **Internalizing difficulties score^c^** | 2.9 (2.7) | 3.0 (2.8) | 0.431 | 2.8 (2.5) | 0.189 | 3.0 (2.8) | 0.068 | 2.9 (2.7) | 0.651 |
| **Externalizing difficulties score^c^** | 4.2 (3.4) | 4.1 (3.4) | 0.483 | 4.4 (3.5) | 0.168 | 4.4 (3.3) | 0.109 | 4.4 (3.4) | 0.147 |

Results are numbers (%) for categorical variables and mean (standard deviation) for continuous ones. P-values compare survey non-respondents to respondents and are from Chi-squared test for categorical variables and analysis of variance for continuous ones. Baseline survey: December 2021 – June 2022; thematic survey: September 2022 – January 2023; 1^st^ follow-up: May – July 2023; 1^st^ follow-up: May – July 2024; 1^st^ follow-up: May – July 2025.

^a^ Not collected among 322 participants < 6 years old.

^b^ Not collected among 61 participants < 3 years old.

^c^ Mental health difficulties assessed with the strengths and difficulties questionnaire (SDQ), with higher values indicating heightened difficulties.

**Additional table 2.** Yearly incidence rate ratio of mental health difficulties according to the COVID-19 pandemic impact, by age category and impact measure in children aged 2-17 years old, between 2022 and 2025 (n=1907).

| **Age category** | **COVID-19 pandemic impact measure** | **Type of impact** | **Mental health difficulties** | | **Internalizing difficulties** | | | **Externalizing difficulties** | |
| --- | --- | --- | --- | --- | --- | --- | --- | --- | --- |
|  |  |  | **Maximum likelihood** | **Multiple imputation** | | **Maximum likelihood** | **Multiple imputation** | **Maximum likelihood** | **Multiple imputation** |
|  |  |  | IRR (95% CI) | IRR (95% CI) | | IRR (95% CI) | IRR (95% CI) | IRR (95% CI) | IRR (95% CI) |
| All | Child impact | None/minimal | Ref. | Ref. | | Ref. | Ref. | Ref. | Ref. |
| All | Child impact | Positive | 1.03 (0.99; 1.07) | 1.02 (0.98; 1.06) | | 1.02 (0.97; 1.09) | 1.02 (0.96; 1.07) | 1.03 (0.98; 1.08) | 1.02 (0.97; 1.06) |
| All | Child impact | Negative | 0.97 (0.94; 1.00)° | 0.97 (0.95; 1.01) | | 0.98 (0.94; 1.02) | 0.97 (0.93; 1.01) | 0.97 (0.93; 1.01)° | 0.98 (0.95; 1.01) |
| All | Household impact | None/minimal | Ref. | Ref. | | Ref. | Ref. | Ref. | Ref. |
| All | Household impact | Positive | 1.02 (0.98; 1.05) | 1.01 (0.98; 1.05) | | 1.01 (0.96; 1.06) | 1.01 (0.96; 1.05) | 1.02 (0.98; 1.07) | 1.02 (0.98; 1.06) |
| All | Household impact | Negative | 0.97 (0.93; 1.00)° | 0.97 (0.94; 1.01) | | 0.98 (0.93; 1.03) | 0.97 (0.92; 1.02) | 0.96 (0.92; 1.00)° | 0.97 (0.93; 1.01) |
| 2-6 | Child impact | None/minimal | Ref. | Ref. | | Ref. | Ref. | Ref. | Ref. |
| 2-6 | Child impact | Positive | 1.04 (0.96; 1.13) | 1.03 (0.96; 1.11) | | 1.06 (0.93; 1.22) | 1.05 (0.92; 1.19) | 1.04 (0.96; 1.13) | 1.03 (0.95; 1.11) |
| 2-6 | Child impact | Negative | 0.96 (0.90; 1.03) | 0.98 (0.92; 1.04) | | 1.02 (0.91; 1.14) | 1.01 (0.91; 1.12) | 0.93 (0.86; 1.01)° | 0.97 (0.91; 1.02) |
| 7-12 | Child impact | None/minimal | Ref. | Ref. | | Ref. | Ref. | Ref. | Ref. |
| 7-12 | Child impact | Positive | 1.03 (0.98; 1.09) | 1.01 (0.97; 1.07) | | 1.02 (0.94; 1.09) | 1.00 (0.93; 1.08) | 1.04 (0.97; 1.11) | 1.02 (0.96; 1.08) |
| 7-12 | Child impact | Negative | 0.97 (0.93; 1.01) | 0.97 (0.93; 1.02) | | 0.95 (0.89; 1.01)° | 0.95 (0.90; 1.01) | 0.98 (0.93; 1.03) | 0.99 (0.94; 1.04) |
| 13-17 | Child impact | None/minimal | Ref. | Ref. | | Ref. | Ref. | Ref. | Ref. |
| 13-17 | Child impact | Positive | 1.02 (0.91; 1.14) | 1.01 (0.92; 1.11) | | 1.04 (0.91; 1.19) | 1.03 (0.92; 1.15) | 0.98 (0.84; 1.15) | 0.99 (0.88; 1.12) |
| 13-17 | Child impact | Negative | 0.99 (0.94; 1.04) | 0.97 (0.92; 1.02) | | 0.99 (0.92; 1.06) | 0.96 (0.90; 1.03) | 0.98 (0.91; 1.05) | 0.98 (0.91; 1.04) |

° p-value < 0.1

Results are incidence rate ratio (IRR) and 95% confidence intervals (CI) of pandemic impact × time interactions from generalized mixed effects models with fixed effects for age, sex, chronic condition, and household financial situation, and random effects at the child and household levels.

**Additional table 3.** Yearly incidence rate ratio of mental health difficulties according to the child COVID‑19 pandemic impact and socio-demographic, health, family, social and behavioral moderators, in children aged 2-17 years old, between 2022 and 2025 (n=1907).

| **Moderator** | **Child**  **COVID-19 pandemic impact** | **Moderator × pandemic** **impact interaction** | | **Moderator × pandemic** **impact × time interaction** | |
| --- | --- | --- | --- | --- | --- |
|  |  | **Maximum likelihood** | **Multiple imputation** | **Maximum likelihood** | **Multiple imputation** |
|  |  | **IRR (95% CI)** | **IRR (95% CI)** | **IRR (95% CI)** | **IRR (95% CI)** |
| **Age category** |  |  |  |  |  |
| 2-6 | None or minimal | Ref. | Ref. | Ref. | Ref. |
| 7-12 | Positive | 1.47 (1.06; 2.02)* | 1.36 (0.98; 1.89)° | 0.99 (0.91; 1.09) | 0.98 (0.89; 1.07) |
| 7-12 | Negative | 0.87 (0.64; 1.19) | 0.90 (0.67; 1.22) | 1.01 (0.93; 1.09) | 0.99 (0.92; 1.07) |
| 13-17 | Positive | 1.26 (0.83; 1.92) | 1.12 (0.76; 1.66) | 0.98 (0.86; 1.12) | 0.98 (0.88; 1.10) |
| 13-17 | Negative | 0.97 (0.69; 1.35) | 1.00 (0.73; 1.36) | 1.03 (0.94; 1.12) | 1.00 (0.92; 1.08) |
| **Sex** |  |  |  |  |  |
| Male | None or minimal | Ref. | Ref. | Ref. | Ref. |
| Female | Positive | 1.07 (0.82; 1.40) | 1.08 (0.84; 1.39) | 0.96 (0.89; 1.04) | 0.98 (0.91; 1.05) |
| Female | Negative | 1.12 (0.89; 1.40) | 1.11 (0.90; 1.37) | 1.04 (0.98; 1.11) | 1.01 (0.96; 1.07) |
| **Chronic condition** |  |  |  |  |  |
| No | None or minimal | Ref. | Ref. | Ref. | Ref. |
| Yes | Positive | 1.23 (0.92; 1.63) | 1.20 (0.89; 1.62) | 1.03 (0.95; 1.12) | 1.00 (0.93; 1.08) |
| Yes | Negative | 1.00 (0.79; 1.25) | 1.00 (0.80; 1.25) | 1.02 (0.96; 1.09) | 1.01 (0.95; 1.07) |
| **Household financial situation** |  |  |  |  |  |
| Very good | None or minimal | Ref. | Ref. | Ref. | Ref. |
| Good | Positive | 0.79 (0.56; 1.12) | 0.84 (0.58; 1.20) | 1.04 (0.95; 1.15) | 1.03 (0.94; 1.13) |
| Good | Negative | 0.73 (0.54; 0.97)* | 0.77 (0.57; 1.04)° | 1.03 (0.96; 1.11) | 1.02 (0.96; 1.09) |
| Average to poor | Positive | 0.91 (0.62; 1.36) | 0.91 (0.61; 1.34) | 1.08 (0.97; 1.19) | 1.04 (0.95; 1.14) |
| Average to poor | Negative | 0.57 (0.40; 0.81)* | 0.64 (0.46; 0.89)* | 1.07 (0.98; 1.17) | 1.05 (0.97; 1.13) |
| **Parents' highest education** |  |  |  |  |  |
| University | None or minimal | Ref. | Ref. | Ref. | Ref. |
| Lower than university | Positive | 0.83 (0.56; 1.23) | 0.90 (0.63; 1.29) | 0.96 (0.86; 1.07) | 0.98 (0.88; 1.08) |
| Lower than university | Negative | 0.84 (0.59; 1.18) | 0.79 (0.58; 1.08) | 0.95 (0.87; 1.05) | 1.00 (0.93; 1.07) |
| **Parent-child relationship** |  |  |  |  |  |
| Very good to good | None or minimal | Ref. | Ref. | Ref. | Ref. |
| Less than good | Positive | 1.17 (0.84; 1.63) | 1.10 (0.80; 1.52) | 1.03 (0.94; 1.13) | 1.01 (0.92; 1.10) |
| Less than good | Negative | 0.83 (0.66; 1.05) | 0.84 (0.67; 1.05) | 1.04 (0.97; 1.10) | 1.03 (0.97; 1.09) |
| **Parents' mood** |  |  |  |  |  |
| Very good to good | None or minimal | Ref. | Ref. | Ref. | Ref. |
| Average to poor | Positive | 1.16 (0.76; 1.76) | 1.21 (0.80; 1.82) | 0.99 (0.89; 1.09) | 0.98 (0.89; 1.08) |
| Average to poor | Negative | 0.76 (0.58; 1.00)° | 0.83 (0.64; 1.08) | 1.06 (0.99; 1.14) | 1.03 (0.97; 1.10) |
| **Number of friends** |  |  |  |  |  |
| Several | None or minimal | Ref. | Ref. | Ref. | Ref. |
| One or less | Positive | 1.00 (0.70; 1.44) | 1.08 (0.75; 1.53) | 0.94 (0.84; 1.05) | 0.97 (0.88; 1.07) |
| One or less | Negative | 1.02 (0.78; 1.34) | 1.09 (0.84; 1.42) | 0.94 (0.88; 1.01)° | 0.97 (0.91; 1.03) |
| **Extracurricular activities** | |  |  |  |  |
| Yes | None or minimal | Ref. | Ref. | Ref. | Ref. |
| No | Positive | 0.75 (0.49; 1.13) | 0.79 (0.53; 1.18) | 0.94 (0.84; 1.05) | 0.98 (0.88; 1.09) |
| No | Negative | 1.11 (0.81; 1.52) | 1.05 (0.77; 1.42) | 0.99 (0.91; 1.07) | 0.99 (0.92; 1.06) |
| **Screen time** |  |  |  |  |  |
| Meeting recommendations | None or minimal | Ref. | Ref. | Ref. | Ref. |
| Not meeting recommendations | Positive | 0.92 (0.65; 1.29) | 0.94 (0.68; 1.29) | 1.04 (0.94; 1.15) | 1.03 (0.94; 1.13) |
| Not meeting recommendations | Negative | 1.02 (0.79; 1.31) | 1.01 (0.79; 1.28) | 0.99 (0.93; 1.06) | 0.99 (0.93; 1.06) |
| **Physical activity** |  |  |  |  |  |
| Meeting recommendations | None or minimal | Ref. | Ref. | Ref. | Ref. |
| Not meeting recommendations | Positive | 0.93 (0.69; 1.25) | 1.00 (0.74; 1.36) | 0.94 (0.86; 1.02) | 0.96 (0.88; 1.05) |
| Not meeting recommendations | Negative | 0.94 (0.74; 1.19) | 0.99 (0.78; 1.26) | 1.00 (0.94; 1.07) | 0.99 (0.94; 1.05) |
| **Sleep duration** |  |  |  |  |  |
| Meeting recommendations | None or minimal | Ref. | Ref. | Ref. | Ref. |
| Not meeting recommendations | Positive | 1.33 (0.93; 1.89) | 1.20 (0.84; 1.71) | 0.96 (0.86; 1.07) | 0.96 (0.87; 1.06) |
| Not meeting recommendations | Negative | 1.05 (0.81; 1.37) | 0.95 (0.75; 1.20) | 0.95 (0.89; 1.02) | 0.98 (0.91; 1.04) |

° p-value < 0.1; * p-value < 0.05. Results are incidence rate ratio (IRR) and 95% confidence intervals (CI) from generalized mixed effects models adjusted with fixed effects for age, sex, chronic condition, and household financial situation, and random effects at the child and household levels.

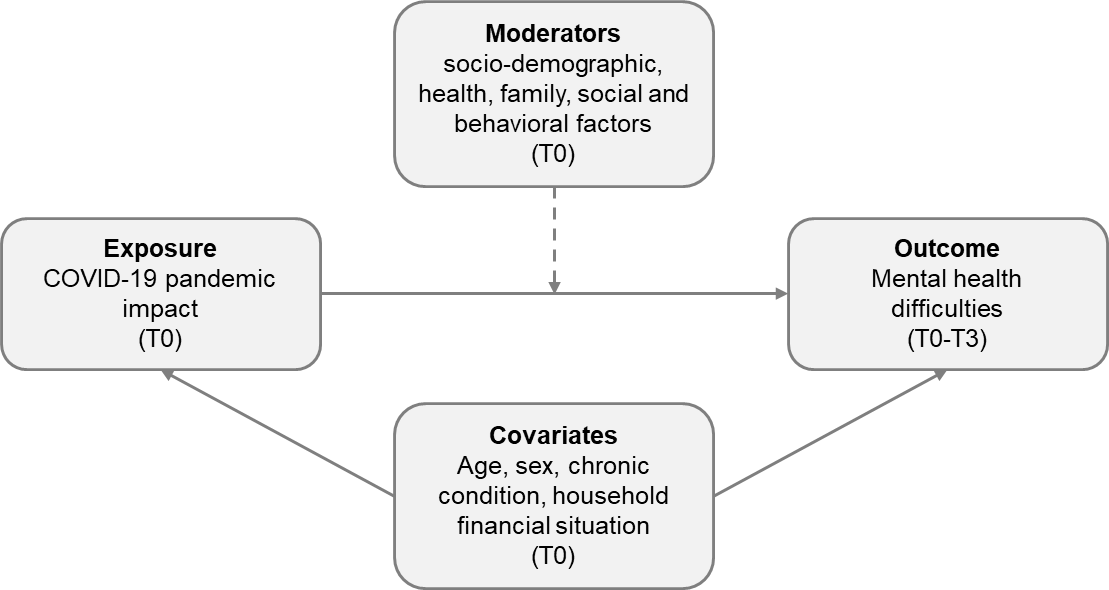

**Additional figure 1.** Directed acyclic graph of the hypothesized relationships underlying the main study model (plain line) and moderator’s models (dashed line). T0 assessments were conducted in 2022, whether in the baseline survey for the mental health and covariates or in a thematic survey for the pandemic impact. T1-T3 assessments were conducted in May-July 2023, 2024, and 2025, respectively.

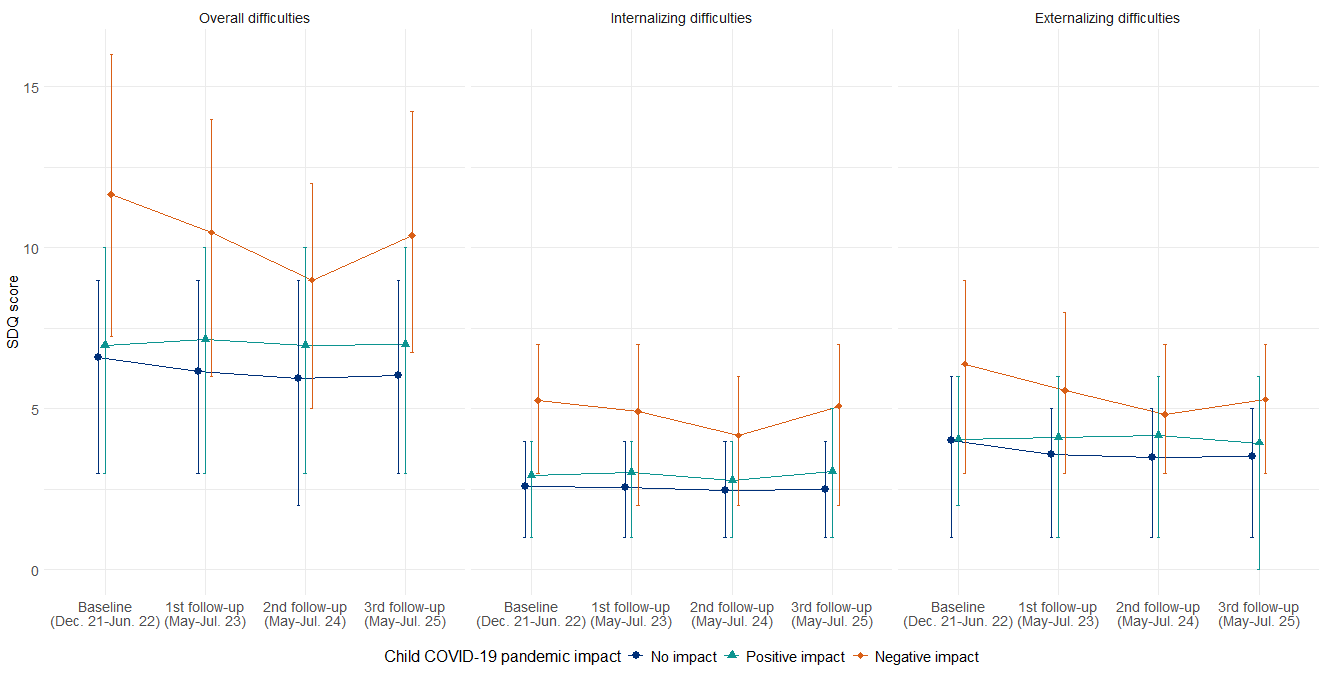

**Additional figure 2.** Median and interquartile range of the strengths and difficulties questionnaire (SDQ) scores across timepoints by child COVID-19 pandemic impact (n=1488).
